# An estimate of pediatric lives saved due to non-pharmacologic interventions during the early COVID-19 pandemic

**DOI:** 10.1101/2023.04.18.23288763

**Authors:** Jeremy Samuel Faust, Benjamin Renton, Chengan Du, Alexander Junxiang Chen, Shu-Xia Li, Zhenqiu Lin, Harlan Krumholz

## Abstract

The net effect of the pandemic mitigation strategies on childhood mortality is not known. During the first year of the COVID-19 pandemic, mitigation policies and behaviors were widespread, and although vaccinations and effective treatments were not yet widely available, the risk of death from SARS-CoV-2 infection was low. In that first year, there was a 7% decrease in medical (“natural causes”) mortality among children ages 0-9 during the first pandemic year (5% among infants <1 year and 15% among children ages 1-9) in the United States, resulting in an estimated 1,488 deaths due to medical causes averted among children ages 0-9, and 1,938 deaths averted over 24 months. The usual expected surge in winter medical deaths, particularly among children ages >1 year was absent. However, smaller increases in external (“non-natural causes”) mortality were also observed during the study period, which decreased the overall number of pediatric deaths averted during both years and the pandemic period. In total, 1,468 fewer all-cause pediatric deaths than expected occurred in the United States during the first 24 months of the COVID-19 pandemic.

## Main

The net effect of the pandemic mitigation strategies on childhood mortality is not known. The mitigation strategies included shelter-in-place, physical distancing, masking, hygiene campaigns, testing, and other measures. During the first year of the COVID-19 pandemic, these mitigation policies and behaviors were widespread, and although vaccinations and effective treatments were not yet widely available, the risk of death from SARS-CoV-2 infection was low in children.

The number of childhood deaths averted during the pandemic is unknown. Mortality from infectious and other diseases decreased during the first year of the pandemic in the United States, but these decreases were eclipsed by COVID-19 deaths, leading to historic all-cause excess mortality in older and younger adults.^1–3^ As a result, in these individuals it is difficult to discern the effect of the mitigation strategies. However, children had very low COVID-19 mortality rates and thus, it is possible to assess the association of pandemic interventions on overall mortality.

Accordingly, as previously (see Supplement), we modeled excess mortality in US children ages 0-9, using monthly mortality (2015-2019) and yearly population data (2014-2019) to project monthly expected medical cause (“natural causes”), external cause (“non-natural causes”) and all-cause deaths and corresponding 95% confidence intervals (CI) for the pandemic starting March 2020.^4,5^ Overall estimates were assembled from component models of smaller age groups (<1, 1-4, 5-9 years). Sensitivity analyses using 3- and 4-year baseline periods for expected deaths were conducted (Figure S1, Table S1-2).

In the first year of pandemic mitigation, 1,302 fewer all-cause deaths (95% CI 614-1,990) than expected occurred among children ages 0-9, a 5% decrease (Table 1). Most of the decreased mortality was detected among medical causes, with a 7% decrease (Figure 1, top row, Figure S2-3, Table 1). Decreased medical mortality during the first year was seen among infants ages <1 year (949 fewer deaths; 95% 542-1,357, a 5% reduction), 1-4-year-olds (351 fewer deaths; 95% 210-493, a 16% reduction), and 5-9-year-olds (187 fewer deaths; 95% 121-254, a 13% reduction) (Table 1). Among non-infants (children ages 1-9-years), 539 (95% CI 381-697) fewer medical deaths were observed during the first 12 months of the pandemic, a 15% reduction.

**Table 1.**

| Period | Field | < 1 years | 1-4 years | 5-9 years | 1-9 years | 0-9 years |
| --- | --- | --- | --- | --- | --- | --- |
| March 2020-February 2021 | All Causes | -970 (-1373 to -567) | -197 (-384 to -10) | -134 (-212 to -56) | -332 (-564 to -100) | -1302 (-1990 to -614) |
| March 2020-February 2021 | Medical | -949 (-1357 to -542) | -351 (-493 to -210) | -187 (-254 to -121) | -539 (-697 to -381) | -1488 (-1956 to -1020) |
| March 2020-February 2021 | External | -21 (-125 to 84) | 154 (37 to 271) | 53 (-17 to 123) | 207 (121 to 293) | 186 (49 to 323) |
| March 2020-February 2021 | COVID UCD (MCD) Deaths | 44 (69) | 25 (32) | 24 (38) | 49 (70) | 93 (139) |
| March 2020-February 2021 | All Causes Observed/Expected Ratio | 0.95 | 0.95 | 0.94 | 0.94 | 0.95 |
| March 2020-February 2021 | Medical Observed/Expected Ratio | 0.95 | 0.84 | 0.87 | 0.85 | 0.93 |
| March 2020-February 2021 | External Observed/Expected Ratio | 0.99 | 1.1 | 1.06 | 1.09 | 1.05 |
| March 2021-February 2022 | All Causes | -305 (-701 to 91) | 15 (-169 to 199) | 123 (48 to 198) | 139 (-89 to 367) | -167 (-852 to 518) |
| March 2021-February 2022 | Medical | -410 (-810 to -9) | -11 (-151 to 129) | -29 (-94 to 36) | -40 (-196 to 115) | -450 (-910 to 11) |
| March 2021-February 2022 | External | 105 (2 to 208) | 26 (-91 to 144) | 152 (83 to 221) | 179 (95 to 262) | 283 (150 to 417) |
| March 2021-February 2022 | COVID UCD (MCD) Deaths | 139 (242) | 75 (107) | 82 (99) | 157 (206) | 296 (448) |
| March 2021-February 2022 | All Causes Observed/Expected Ratio | 0.98 | 1 | 1.05 | 1.02 | 0.99 |
| March 2021-February 2022 | Medical Observed/Expected Ratio | 0.98 | 0.99 | 0.98 | 0.99 | 0.98 |
| March 2021-February 2022 | External Observed/Expected Ratio | 1.07 | 1.02 | 1.17 | 1.07 | 1.07 |
| Pandemic period | All Causes | -1275 (-1980 to -570) | -181 (-497 to 135) | -12 (-141 to 117) | -193 (-622 to 236) | -1468 (-2701 to -235) |
| Pandemic period | Medical | -1359 (-1943 to -775) | -362 (-622 to -103) | -217 (-328 to -106) | -579 (-861 to -297) | -1938 (-2688 to -1188) |
| Pandemic period | External | 84 (-66 to 234) | 181 (-169 to 530) | 205 (72 to 339) | 386 (228 to 544) | 470 (206 to 733) |
| Pandemic period | COVID UCD (MCD) Deaths | 183 (311) | 100 (139) | 106 (137) | 206 (276) | 389 (587) |
| Pandemic period | All Causes Observed/Expected Ratio | 0.97 | 0.98 | 1 | 0.98 | 0.97 |
| Pandemic period | Medical Observed/Expected Ratio | 0.96 | 0.92 | 0.92 | 0.92 | 0.96 |
| Pandemic period | External Observed/Expected Ratio | 1.03 | 1.06 | 1.11 | 1.08 | 1.06 |

**Figure 1.**
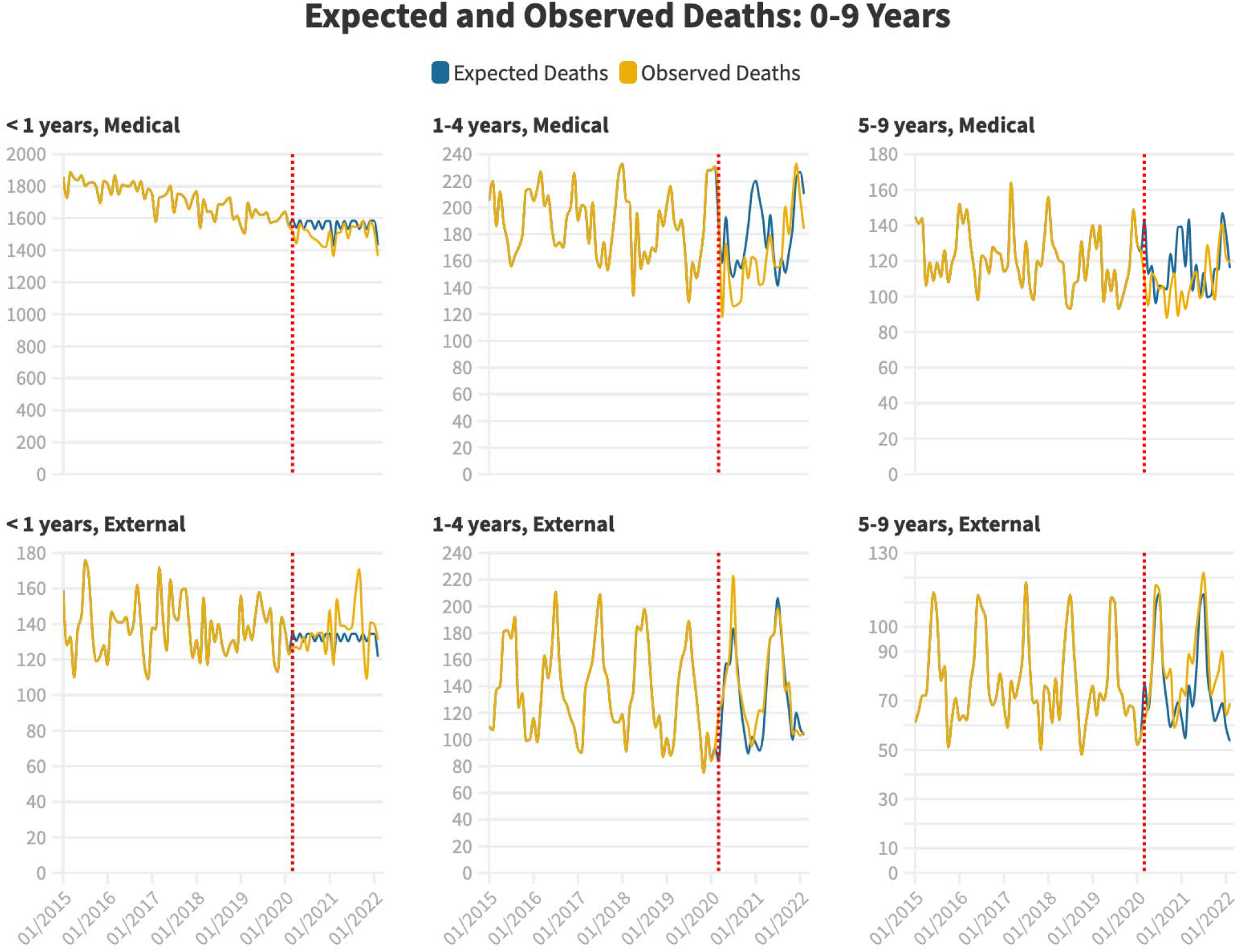
Monthly observed raw deaths are shown in yellow. Modeled expected deaths during the Covid-19 pandemic period are shown in blue. All-cause expected deaths (left panel) include all ICD-10 classifications that appear in CDC WONDER; Medical expected deaths (top row) include all ICD-10 classifications with the exception of ICD chapters V01-Y89; External deaths (bottom row) include ICD chapters V01-Y89.^4^

The greatest relative decreases in all-cause and medical mortality were observed in December 2020 for <1-year-olds (10% and 10%). For 1-4-year-olds, the greatest relative decreases were observed in December 2020 both for all-cause mortality (18%) and medical mortality (11%). For 5-9-year-olds, the greatest relative decreases were observed in December 2020 for all-cause mortality (27%) and March 2020 for medical mortality (19%) (Table S3-5).

Deaths from common infectious diseases were lower than usual, but changes among several typical leading cause of death varied by age (Figure S4). External mortality increased (186 excess deaths; 95% CI 49-323, a 5% increase), driven by older children (Figure 1, bottom row, Table 1, Figure S5). The most common cause of external mortality was accidental injury not involving traffic (Figure S6).

During the second year (when fewer non-pharmacological interventions were in place), decreases in all-cause and medical mortality abated (Figure 1, top row, Table 1), though excess external mortality occurred (283 excess deaths; 95% CI 150-417) (Figure 1, bottom row). Pediatric Covid-19-specific mortality also increased during this period (Table 1).

Overall, there were 1,468 fewer all-cause deaths (95% CI 235-2,701), 1,938 fewer medical deaths (95% CI 1,188-2,688), and 470 more external cause deaths (95% CI 206-733) among US children ages 0-9 during the first 2 pandemic years (Figure 2, Table 1).

**Figure 2.**
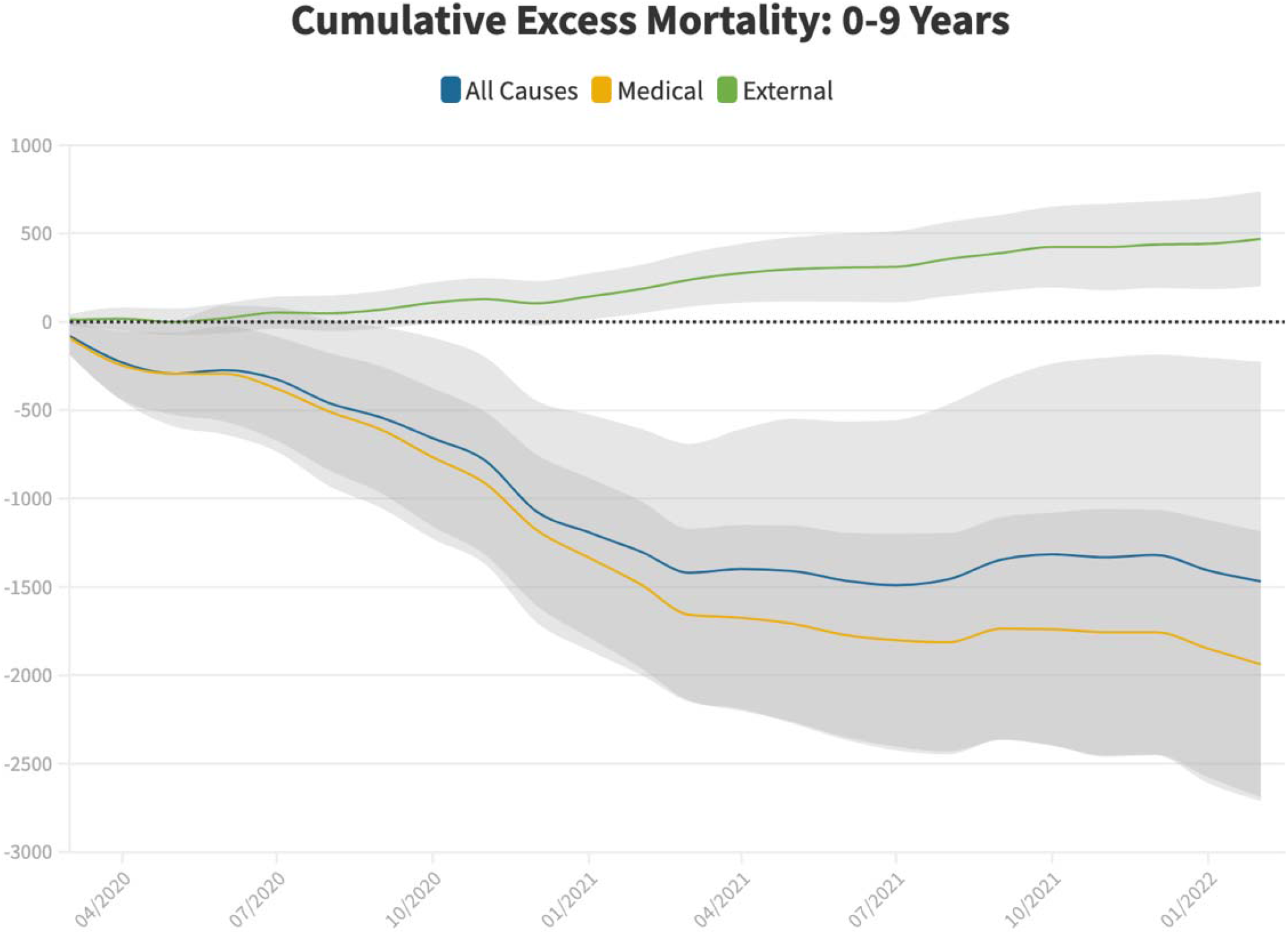
Cumulative raw excess deaths (monthly data) are shown for all-causes (blue line), medical causes (yellow line), and external causes (green line). All-cause excess deaths include all ICD-10 classifications that appear in CDC WONDER; Medical excess deaths include all ICD-10 classifications with the exception of ICD chapters V01-Y89; External excess deaths include ICD chapters V01-Y89.^4^ 95% confidence intervals are indicated by the gray shaded regions. Sensitivity analyses using 3- and 4-year baseline periods to generate expected deaths are shown in Figure S1 and summarized in Table S1-2.

Modeling and historical data indicate that each winter there is an expected surge in medical deaths among children ages >1 year, similar to patterns known to occur among adults.^6^ Covid-19-related mitigation clearly disrupted and eliminated the expected annual winter surge in medical deaths during the first full pandemic winter (December 2020-March 2021), as shown in Figure 1 (see the top middle and top right panels; note the area between the modeled number of expected deaths shown in blue and raw observed deaths, shown in yellow, and the absence of a spike in medical mortality that routinely seen in all pre-pandemic years shown to the left of the red dashed line).

Historical data also show that external deaths in this age group typically increase in the summer. The present data show that unlike medical deaths, the typical seasonal trends in external mortality were not disrupted by pandemic-related behavioral changes (see bottom middle and bottom right panels, in which expected deaths shown in blue and observed deaths shown in yellow continued to track closely) but were nonetheless statistically higher than expected.

This natural experiment indicates that pandemic interventions likely averted thousands of pediatric deaths due to medical causes, most notably during the first full winter (2021), when interventions were most intense across the nation and the virus had not yet reached all parts of the country. In that winter of 2021, the usual seasonal increase in medical causes of childhood mortality was effectively eliminated.

## Methods

We conducted our analysis with R version 4.0.3. Studies of public data are exempt from institutional review approval (HHS regulation 45-CFR-46.101(c)).

To project expected deaths, we used seasonal autoregressive integrated moving averages from January 2015 up to February 2020.

We assume there were 12 months within year 2020. For each month, we defined the original projected population for five-year age groups (ages <1, 1-4, 5-9), (denoted by *k*, where 1 ≤ *k* ≤ 4) in month *i* of 2020 to be *n*_*i,k*_; the original point estimate for all-cause deaths to be 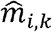; the sum of all previous months’ excess death to be *P*_*i-*1,*k*_ (starting from March 2020). Therefore, the corrected expected deaths for month *i* (since March 2020) after correction 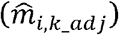 satisfies:

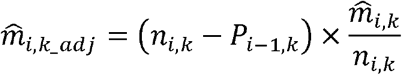

The above equation also holds for the months in 2021 and 2022, where *P*_*i-*1,*k*_ denotes the sum of all previous months’ excess death since March 2020, respectively.

The sARIMA models in the study were based on monthly data and set the month indicators as well as the estimated monthly population as covariates. By adding the month indicators, we can not only capture the seasonal trends well in the models but also increase our model predictability by considering the month effects. Moreover, by adding the population, we can capture the death counts dynamically influenced by the number of people in each group.

The models in the study were given the death counts for the three age groups (ages <1, 1-4, 5-9) and the total for each state for all adults from January 2015, up to February 2020, a total of 62 months previous to COVID-19, to ensure that our projections were based both on smooth data trends and sufficient data. Composite point estimates summed from component models for expected deaths are rounded to the nearest whole number.

The monthly 95% CI boundaries directly from the sARIMA model in the auto.arima function in the R statistical software. The periodically CI boundaries were obtained through the 5,000 simulation samples from the estimated sARIMA model for each age group.

All-cause deaths were divided into medical manner of death and external manner of death. Medical manner causes of death are divided into 19 ICD chapters, which are: A00-B99 (Certain infectious and parasitic diseases), C00-D48 (Neoplasms), D50-D89 (Diseases of the blood and blood-forming organs and certain disorders involving the immune mechanism), E00-E88 (Endocrine, nutritional and metabolic diseases), F01-F99 (Mental and behavioral disorders), G00-G98 (Diseases of the nervous system), H00-H57 (Diseases of the eye and adnexa), H60-H93 (Diseases of the ear and mastoid process), I00-I99 (Diseases of the circulatory system), J00-J98 (Diseases of the respiratory system), K00-K92 (Diseases of the digestive system), L00-L98 (Diseases of the skin and subcutaneous tissue), M00-M99 (Diseases of the musculoskeletal system and connective tissue), N00-N98 (Diseases of the genitourinary system), O00-O99 (Certain conditions originating in the perinatal period), Q00-Q99 (Congenital malformations, deformations and chromosomal abnormalities), R00-R99 (Symptoms signs and abnormal clinical and laboratory findings, not elsewhere classified) and U00-U99 (Codes for special purposes [which includes COVID-19]).

External manner causes of death are under the ICD chapters V01-Y89, which are subdivided into V01-V99 (Transport accidents), W00-59 (Other external causes of accidental injury), X60-X84 (Intentional self-harm), X85-Y09 (Assault), Y10-Y34 (Event of undetermined intent), Y35-Y36 (Legal intervention and operations of war), Y40-Y84 (Complications of medical and surgical care) and Y85-Y89 (Sequelae of external causes of morbidity and mortality).

## Data Availability

All data produced in the present study are available upon reasonable request to the authors

## Disclosures

Conflict of Interest Disclosures: Dr. Krumholz reported receiving consulting fees from UnitedHealth, Element Science, Aetna, Reality Labs, F-Prime, and Tesseract/4Catalyst; serving as an expert witness for Martin/Baughman law firm, Arnold and Porter law firm, and Siegfried and Jensen law firm; being a cofounder of Hugo Health, a personal health information platform; being a cofounder of Refactor Health, an enterprise health care, artificial intelligence–augmented data management company; receiving contracts from the Centers for Medicare & Medicaid Services through Yale New Haven Hospital to develop and maintain performance measures that are publicly reported; and receiving grants from Johnson & Johnson outside the submitted work. No other disclosures were reported.

## Notes

### Funding Statement

This study did not receive any funding

