## Supplemental Appendix for "An estimate of pediatric lives saved due to non-pharmacologic interventions during the early COVID-19 pandemic"

#### **Supplemental Exhibits List**

Figure 1. Monthly expected and observed deaths, all, medical and external causes, Ages 0-9 years, 3- and 4-year baseline periods for modeling, January 2015-February 2022.

Table 1. Excess deaths, all, medical and external causes, and COVID-19 deaths, March 2020-February 2022, modeled using 4-year baseline period.

Table 2. Excess deaths, all, medical and external causes, and COVID-19 deaths, March 2020-February 2022, modeled using 3-year baseline period.

Figure 2. Monthly expected and observed deaths, all, medical and external causes, Ages 0-9 years, January 2015-February 2022.

Figure 3. Monthly expected and observed deaths, all, medical and external causes, Ages <1, 1-4, 5-9 years, January 2015-February 2022.

Table 3. Excess deaths, all, medical and external causes, by month, Ages < 1 years, March 2020-February 2022.

Table 4. Excess deaths, all, medical and external causes, by month, Ages 1-4 years, March 2020-February 2022.

Table 5. Excess deaths, all, medical and external causes, by month, Ages 5-9 years, March 2020-February 2022.

Figure 4. Deaths by ICD-10 chapter, 12-month periods beginning each March, ages <1 years, March 2010-February 2022.

Figure 5. Deaths by ICD-10 chapter, 12-month periods beginning each March, ages 1-9 years, March 2010-February 2022.

Figure 6. Deaths from external causes by ICD-10 Sub-chapter, ages 0-9 years, March 2010-February 2022 (select causes not shown due to suppression constraints).

#### Supplemental Figure 1

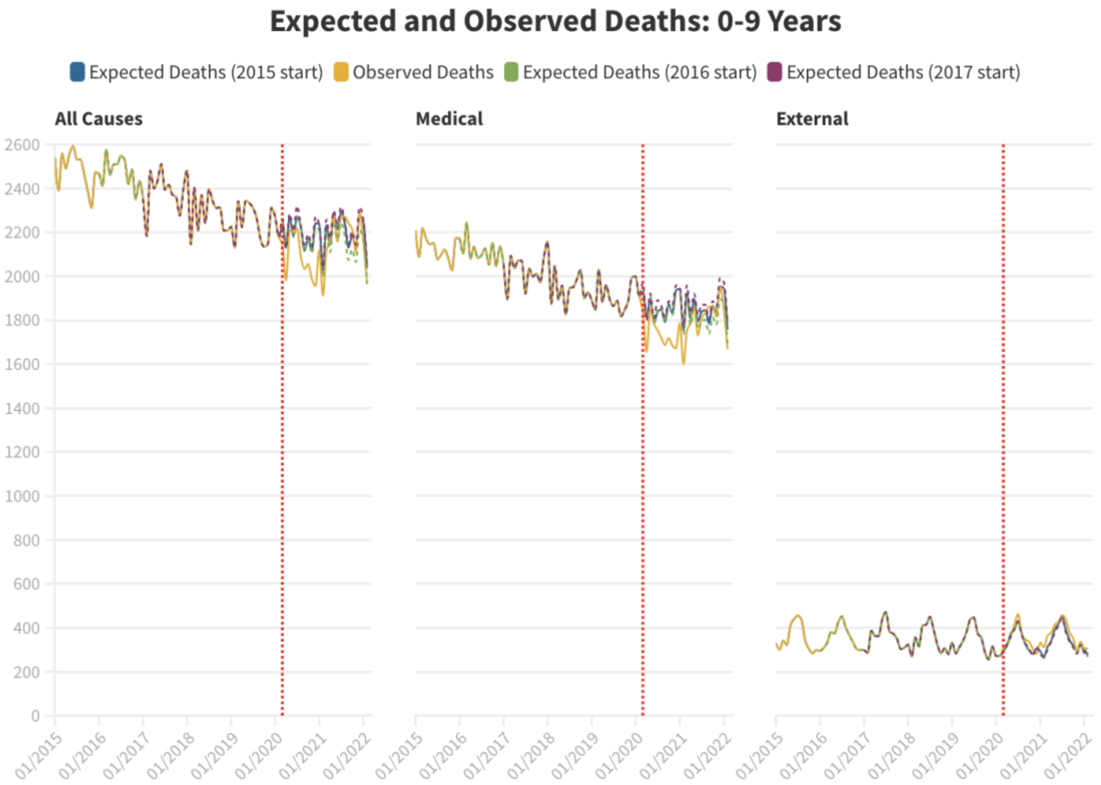

### Supplemental Table 1

| Period | Field | < 1 years | 1-4 years | 5-9 years | 1-9 years | 0-9 years |
|---|---|---|---|---|---|---|
| March 2020-February 2021 | All Causes | -936 (-1360 to -512) | -187 (-366 to -8) | -146 (-227 to -65) | -334 (-542 to -126) | -1270 (-3808 to 1268) |
| March 2020-February 2021 | Medical | -913 (-1315 to -511) | -374 (-573 to -176) | -195 (-257 to -134) | -570 (-784 to -356) | -1483 (-1977 to -989) |
| March 2020-February 2021 | External | -23 (-120 to 74) | 187 (121 to 253) | 49 (-21 to 119) | 236 (152 to 319) | 213 (100 to 327) |
| March 2020-February 2021 | COVID UCD (MCD) Deaths | 44 (69) | 25 (32) | 24 (38) | 49 (70) | 93 (139) |
| March 2020-February 2021 | All Causes Observed/Expected Ratio | 0.95 | 0.95 | 0.94 | 0.94 | 0.95 |
| March 2020-February 2021 | Medical Observed/Expected Ratio | 0.95 | 0.83 | 0.86 | 0.84 | 0.93 |
| March 2020-February 2021 | External Observed/Expected Ratio | 0.99 | 1.13 | 1.05 | 1.1 | 1.05 |
| March 2021-February 2022 | All Causes | 252 (-165 to 669) | 105 (-71 to 281) | 115 (36 to 194) | 220 (16 to 424) | 472 (-2082 to 3026) |
| March 2021-February 2022 | Medical | 129 (-267 to 525) | -15 (-213 to 183) | -33 (-93 to 26) | -48 (-259 to 163) | 81 (-405 to 566) |
| March 2021-February 2022 | External | 123 (27 to 218) | 120 (54 to 185) | 148 (80 to 217) | 268 (185 to 351) | 391 (280 to 502) |
| March 2021-February 2022 | COVID UCD (MCD) Deaths | 139 (242) | 75 (107) | 82 (99) | 157 (206) | 296 (448) |
| March 2021-February 2022 | All Causes Observed/Expected Ratio | 1.01 | 1.03 | 1.05 | 1.04 | 1.02 |
| March 2021-February 2022 | Medical Observed/Expected Ratio | 1.01 | 0.99 | 0.98 | 0.99 | 1 |
| March 2021-February 2022 | External Observed/Expected Ratio | 1.08 | 1.08 | 1.16 | 1.11 | 1.1 |
| Pandemic period | All Causes | -684 (-1292 to -76) | -82 (-398 to 234) | -32 (-176 to 112) | -114 (-435 to 207) | -798 (-8315 to 6719) |
| Pandemic period | Medical | -784 (-1361 to -207) | -389 (-753 to -26) | -229 (-326 to -132) | -618 (-1005 to -231) | -1402 (-2110 to -694) |
| Pandemic period | External | 100 (-39 to 239) | 307 (185 to 429) | 197 (66 to 328) | 504 (350 to 659) | 604 (401 to 808) |
| Pandemic period | COVID UCD (MCD) Deaths | 183 (311) | 100 (139) | 106 (137) | 206 (276) | 389 (587) |
| Pandemic period | All Causes Observed/Expected Ratio | 0.98 | 0.99 | 0.99 | 0.99 | 0.98 |
| Pandemic period | Medical Observed/Expected Ratio | 0.98 | 0.91 | 0.92 | 0.91 | 0.97 |
| Pandemic period | External Observed/Expected Ratio | 1.03 | 1.1 | 1.11 | 1.1 | 1.08 |

### Supplemental Table 2

| Period | Field | < 1 years | 1-4 years | 5-9 years | 1-9 years | 0-9 years |
|---|---|---|---|---|---|---|
| March 2020-February 2021 | All Causes | -1219 (-1662 to -776) | -172 (-331 to -13) | -156 (-219 to -93) | -329 (-576 to -82) | -1548 (-1994 to -1102) |
| March 2020-February 2021 | Medical | -1182 (-1596 to -768) | -277 (-369 to -185) | -213 (-286 to -140) | -490 (-644 to -336) | -1672 (-2164 to -1180) |
| March 2020-February 2021 | External | -37 (-132 to 58) | 105 (34 to 175) | 57 (-11 to 124) | 161 (42 to 281) | 124 (8 to 240) |
| March 2020-February 2021 | COVID UCD (MCD) Deaths | 44 (69) | 25 (32) | 24 (38) | 49 (70) | 93 (139) |
| March 2020-February 2021 | All Causes Observed/Expected Ratio | 0.94 | 0.95 | 0.93 | 0.94 | 0.94 |
| March 2020-February 2021 | Medical Observed/Expected Ratio | 0.94 | 0.87 | 0.85 | 0.86 | 0.93 |
| March 2020-February 2021 | External Observed/Expected Ratio | 0.98 | 1.07 | 1.06 | 1.07 | 1.03 |
| March 2021-February 2022 | All Causes | -621 (-1057 to -185) | 190 (34 to 346) | 116 (54 to 178) | 306 (63 to 549) | -316 (-755 to 123) |
| March 2021-February 2022 | Medical | -710 (-1117 to -303) | 243 (152 to 333) | -34 (-105 to 37) | 209 (57 to 360) | -501 (-985 to -17) |
| March 2021-February 2022 | External | 89 (-5 to 182) | -53 (-123 to 17) | 150 (83 to 216) | 97 (-21 to 214) | 185 (72 to 299) |
| March 2021-February 2022 | COVID UCD (MCD) Deaths | 139 (242) | 75 (107) | 82 (99) | 157 (206) | 296 (448) |
| March 2021-February 2022 | All Causes Observed/Expected Ratio | 0.97 | 1.05 | 1.05 | 1.05 | 0.99 |
| March 2021-February 2022 | Medical Observed/Expected Ratio | 0.96 | 1.13 | 0.98 | 1.06 | 0.98 |
| March 2021-February 2022 | External Observed/Expected Ratio | 1.06 | 0.97 | 1.16 | 1.04 | 1.04 |
| Pandemic period | All Causes | -1841 (-2477 to -1205) | 18 (-239 to 275) | -41 (-172 to 90) | -23 (-454 to 408) | -1864 (-2581 to -1147) |
| Pandemic period | Medical | -1892 (-2485 to -1298) | -34 (-177 to 108) | -247 (-404 to -90) | -281 (-534 to -29) | -2173 (-2879 to -1467) |
| Pandemic period | External | 51 (-85 to 187) | 52 (-103 to 207) | 206 (55 to 358) | 258 (51 to 465) | 309 (104 to 515) |
| Pandemic period | COVID UCD (MCD) Deaths | 183 (311) | 100 (139) | 106 (137) | 206 (276) | 389 (587) |
| Pandemic period | All Causes Observed/Expected Ratio | 0.96 | 1 | 0.99 | 1 | 0.96 |
| Pandemic period | Medical Observed/Expected Ratio | 0.95 | 0.99 | 0.91 | 0.96 | 0.95 |
| Pandemic period | External Observed/Expected Ratio | 1.02 | 1.02 | 1.11 | 1.05 | 1.04 |

#### Supplemental Figure 2

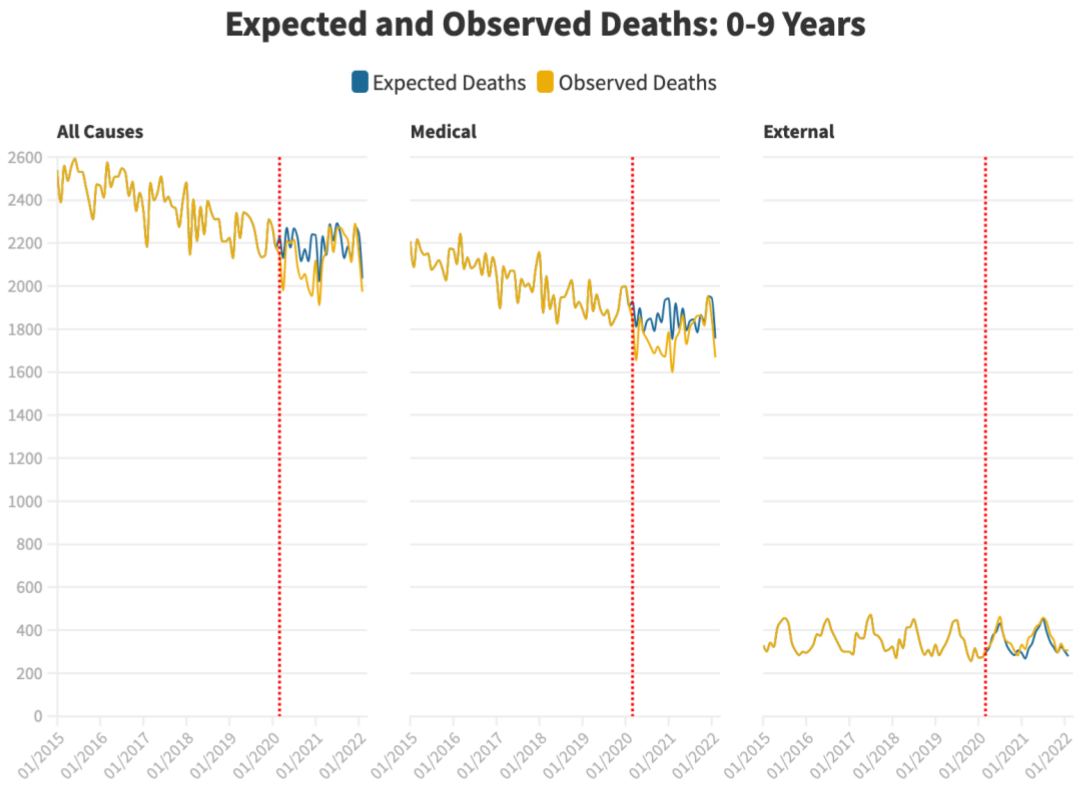

**Supplemental Figure 3**

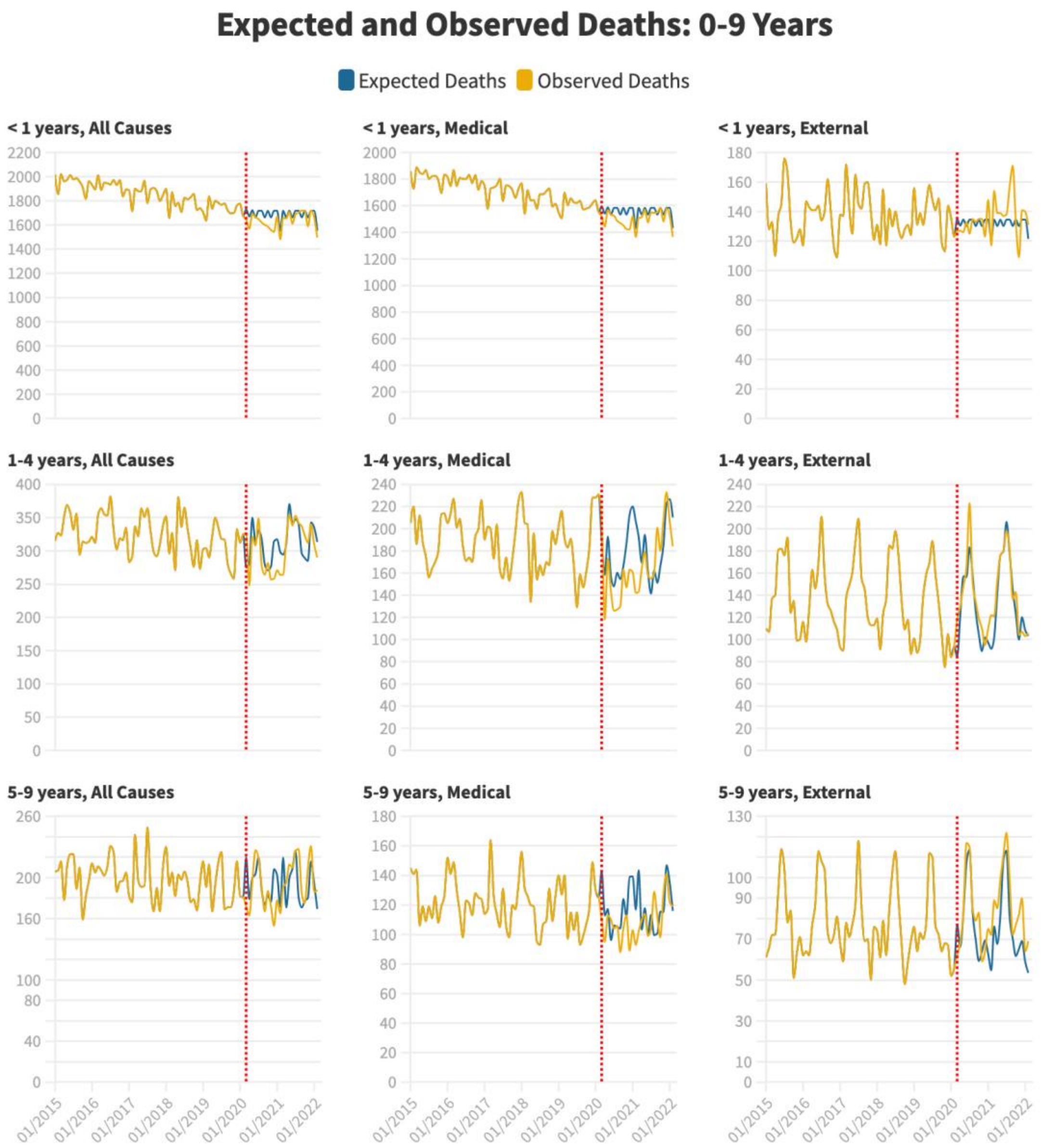

### Supplemental Table 3

| Month | Age group | Excess Deaths (95%) CI | | | Ratio of Observed/Expected (95% CI) | | |
|---|---|---|---|---|---|---|---|
| | | All Causes | Medical | External | All Causes | Medical | External |
| 202003 | < 1 years | -85 (-166 - -3) | -77 (-159 - 6) | -8 (-29 - 13) | 0.95 (0.91 - 1.00) | 0.95 (0.90 - 1.00) | 0.94 (0.81 - 1.12) |
| 202004 | < 1 years | -102 (-183 - -21) | -98 (-181 - -16) | -3 (-25 - 18) | 0.94 (0.90 - 0.99) | 0.94 (0.89 - 0.99) | 0.97 (0.84 - 1.16) |
| 202005 | < 1 years | -30 (-113 - 52) | -22 (-106 - 62) | -9 (-30 - 13) | 0.98 (0.94 - 1.03) | 0.99 (0.94 - 1.04) | 0.94 (0.81 - 1.11) |
| 202006 | < 1 years | 1 (-81 - 84) | -1 (-85 - 82) | 3 (-19 - 24) | 1.00 (0.95 - 1.05) | 1.00 (0.95 - 1.06) | 1.02 (0.88 - 1.22) |
| 202007 | < 1 years | -69 (-149 - 11) | -59 (-141 - 22) | -10 (-30 - 11) | 0.96 (0.92 - 1.01) | 0.96 (0.92 - 1.01) | 0.93 (0.80 - 1.10) |
| 202008 | < 1 years | -97 (-179 - -15) | -98 (-180 - -15) | 0 (-21 - 22) | 0.94 (0.90 - 0.99) | 0.94 (0.89 - 0.99) | 1.00 (0.87 - 1.19) |
| 202009 | < 1 years | -61 (-142 - 20) | -63 (-146 - 19) | 3 (-18 - 24) | 0.96 (0.92 - 1.01) | 0.96 (0.91 - 1.01) | 1.02 (0.88 - 1.22) |
| 202010 | < 1 years | -131 (-214 - -48) | -132 (-216 - -47) | 0 (-21 - 22) | 0.92 (0.88 - 0.97) | 0.92 (0.87 - 0.97) | 1.00 (0.86 - 1.20) |
| 202011 | < 1 years | -104 (-186 - -22) | -109 (-192 - -25) | 5 (-17 - 26) | 0.94 (0.89 - 0.99) | 0.93 (0.88 - 0.98) | 1.04 (0.89 - 1.24) |
| 202012 | < 1 years | -175 (-258 - -92) | -164 (-248 - -79) | -12 (-33 - 10) | 0.90 (0.86 - 0.94) | 0.90 (0.85 - 0.95) | 0.91 (0.79 - 1.09) |
| 202101 | < 1 years | -48 (-129 - 33) | -62 (-144 - 20) | 13 (-8 - 35) | 0.97 (0.93 - 1.02) | 0.96 (0.91 - 1.01) | 1.10 (0.95 - 1.30) |
| 202102 | < 1 years | -70 (-152 - 12) | -66 (-148 - 17) | -5 (-26 - 17) | 0.95 (0.91 - 1.01) | 0.95 (0.90 - 1.01) | 0.96 (0.82 - 1.17) |
| 202103 | < 1 years | -58 (-140 - 23) | -78 (-160 - 5) | 19 (-2 - 41) | 0.97 (0.92 - 1.01) | 0.95 (0.90 - 1.00) | 1.14 (0.99 - 1.36) |
| 202104 | < 1 years | -10 (-91 - 71) | -19 (-101 - 63) | 9 (-12 - 30) | 0.99 (0.95 - 1.04) | 0.99 (0.94 - 1.04) | 1.07 (0.92 - 1.27) |
| 202105 | < 1 years | -9 (-93 - 74) | -14 (-99 - 71) | 4 (-17 - 26) | 0.99 (0.95 - 1.05) | 0.99 (0.94 - 1.05) | 1.03 (0.89 - 1.23) |
| 202106 | < 1 years | -53 (-134 - 28) | -60 (-142 - 22) | 7 (-14 - 28) | 0.97 (0.92 - 1.02) | 0.96 (0.91 - 1.02) | 1.05 (0.91 - 1.26) |
| 202107 | < 1 years | -30 (-111 - 50) | -34 (-116 - 48) | 3 (-18 - 25) | 0.98 (0.94 - 1.03) | 0.98 (0.93 - 1.03) | 1.03 (0.89 - 1.22) |
| 202108 | < 1 years | -16 (-98 - 66) | -39 (-122 - 44) | 22 (1 - 44) | 0.99 (0.95 - 1.04) | 0.98 (0.93 - 1.03) | 1.17 (1.01 - 1.39) |
| 202109 | < 1 years | 57 (-24 - 138) | 16 (-66 - 98) | 41 (20 - 62) | 1.03 (0.99 - 1.09) | 1.01 (0.96 - 1.07) | 1.31 (1.13 - 1.57) |
| 202110 | < 1 years | -1 (-83 - 81) | 0 (-83 - 83) | -2 (-23 - 20) | 1.00 (0.95 - 1.05) | 1.00 (0.95 - 1.06) | 0.99 (0.85 - 1.18) |
| 202111 | < 1 years | -74 (-155 - 8) | -53 (-136 - 30) | -21 (-43 - 0) | 0.96 (0.91 - 1.00) | 0.97 (0.92 - 1.02) | 0.84 (0.72 - 1.00) |
| 202112 | < 1 years | 2 (-79 - 82) | -5 (-87 - 77) | 6 (-15 - 27) | 1.00 (0.96 - 1.05) | 1.00 (0.95 - 1.05) | 1.05 (0.91 - 1.24) |
| 202201 | < 1 years | -54 (-136 - 27) | -60 (-143 - 23) | 5 (-16 - 27) | 0.97 (0.92 - 1.02) | 0.96 (0.91 - 1.02) | 1.04 (0.90 - 1.24) |
| 202202 | < 1 years | -56 (-138 - 26) | -66 (-149 - 18) | 9 (-12 - 31) | 0.96 (0.92 - 1.02) | 0.95 (0.90 - 1.01) | 1.08 (0.92 - 1.31) |

### Supplemental Table 4

| Month | Age group | Excess Deaths (95%) CI | | | Ratio of Observed/Expected (95% CI) | | |
|---|---|---|---|---|---|---|---|
| | | All Causes | Medical | External | All Causes | Medical | External |
| 202003 | 1-4 years | 52 (16 - 89) | 34 (18 - 50) | 18 (-7 - 43) | 1.19 (1.05 - 1.37) | 1.40 (1.18 - 1.73) | 1.10 (0.97 - 1.26) |
| 202004 | 1-4 years | -33 (-69 - 3) | 8 (-7 - 23) | -40 (-65 - -16) | 0.88 (0.78 - 1.01) | 1.06 (0.95 - 1.21) | 0.74 (0.64 - 0.88) |
| 202005 | 1-4 years | -29 (-66 - 8) | -9 (-25 - 6) | -20 (-45 - 6) | 0.92 (0.83 - 1.03) | 0.94 (0.86 - 1.05) | 0.90 (0.79 - 1.03) |
| 202006 | 1-4 years | -5 (-42 - 32) | 9 (-7 - 24) | -14 (-39 - 12) | 0.98 (0.88 - 1.12) | 1.06 (0.96 - 1.17) | 0.91 (0.79 - 1.09) |
| 202007 | 1-4 years | 18 (-18 - 54) | 40 (24 - 55) | -22 (-47 - 3) | 1.05 (0.95 - 1.18) | 1.22 (1.12 - 1.33) | 0.85 (0.73 - 1.02) |
| 202008 | 1-4 years | -40 (-77 - -3) | -7 (-22 - 9) | -33 (-59 - -8) | 0.87 (0.78 - 0.99) | 0.96 (0.87 - 1.06) | 0.79 (0.68 - 0.94) |
| 202009 | 1-4 years | -15 (-52 - 21) | 9 (-6 - 24) | -25 (-49 - 0) | 0.94 (0.84 - 1.09) | 1.07 (0.96 - 1.22) | 0.84 (0.72 - 1.00) |
| 202010 | 1-4 years | 11 (-26 - 48) | 14 (-2 - 30) | -3 (-29 - 22) | 1.04 (0.92 - 1.21) | 1.14 (0.99 - 1.34) | 0.98 (0.85 - 1.16) |
| 202011 | 1-4 years | -19 (-56 - 17) | 21 (5 - 36) | -40 (-65 - -15) | 0.93 (0.82 - 1.07) | 1.23 (1.05 - 1.49) | 0.79 (0.69 - 0.91) |
| 202012 | 1-4 years | -56 (-94 - -19) | -7 (-22 - 9) | -49 (-75 - -24) | 0.82 (0.73 - 0.93) | 0.93 (0.81 - 1.10) | 0.77 (0.69 - 0.87) |
| 202101 | 1-4 years | -46 (-83 - -10) | 13 (-3 - 28) | -59 (-84 - -34) | 0.85 (0.77 - 0.96) | 1.13 (0.98 - 1.34) | 0.73 (0.66 - 0.82) |
| 202102 | 1-4 years | -34 (-71 - 3) | 30 (15 - 46) | -64 (-90 - -39) | 0.89 (0.79 - 1.01) | 1.33 (1.14 - 1.60) | 0.69 (0.61 - 0.78) |
| 202103 | 1-4 years | -31 (-67 - 6) | 19 (4 - 35) | -50 (-75 - -25) | 0.90 (0.80 - 1.02) | 1.19 (1.04 - 1.40) | 0.74 (0.66 - 0.85) |
| 202104 | 1-4 years | 6 (-30 - 42) | 12 (-4 - 27) | -5 (-30 - 19) | 1.02 (0.91 - 1.15) | 1.08 (0.97 - 1.21) | 0.97 (0.85 - 1.13) |
| 202105 | 1-4 years | -16 (-53 - 22) | 0 (-16 - 16) | -16 (-41 - 10) | 0.96 (0.87 - 1.07) | 1.00 (0.92 - 1.10) | 0.92 (0.81 - 1.06) |
| 202106 | 1-4 years | -3 (-39 - 34) | 0 (-15 - 16) | -3 (-28 - 22) | 0.99 (0.90 - 1.11) | 1.00 (0.92 - 1.10) | 0.98 (0.85 - 1.16) |
| 202107 | 1-4 years | 5 (-31 - 42) | -8 (-24 - 7) | 14 (-11 - 38) | 1.02 (0.92 - 1.13) | 0.96 (0.89 - 1.04) | 1.10 (0.93 - 1.33) |
| 202108 | 1-4 years | 2 (-34 - 38) | 4 (-11 - 20) | -2 (-27 - 23) | 1.01 (0.91 - 1.13) | 1.02 (0.94 - 1.12) | 0.99 (0.85 - 1.17) |
| 202109 | 1-4 years | 41 (5 - 77) | -9 (-24 - 6) | 50 (25 - 75) | 1.14 (1.01 - 1.30) | 0.94 (0.85 - 1.05) | 1.33 (1.14 - 1.59) |
| 202110 | 1-4 years | 34 (-3 - 70) | 20 (4 - 36) | 14 (-11 - 39) | 1.12 (0.99 - 1.28) | 1.16 (1.03 - 1.33) | 1.08 (0.94 - 1.28) |
| 202111 | 1-4 years | 27 (-9 - 64) | 4 (-12 - 20) | 23 (-2 - 48) | 1.10 (0.97 - 1.26) | 1.04 (0.90 - 1.24) | 1.13 (0.99 - 1.30) |
| 202112 | 1-4 years | -3 (-39 - 33) | -13 (-29 - 2) | 10 (-14 - 35) | 0.99 (0.90 - 1.11) | 0.89 (0.79 - 1.02) | 1.05 (0.94 - 1.18) |
| 202201 | 1-4 years | -25 (-62 - 12) | -6 (-21 - 10) | -20 (-45 - 6) | 0.92 (0.83 - 1.04) | 0.95 (0.83 - 1.11) | 0.91 (0.82 - 1.03) |
| 202202 | 1-4 years | -23 (-60 - 13) | 3 (-13 - 18) | -26 (-51 - -1) | 0.93 (0.83 - 1.05) | 1.03 (0.89 - 1.21) | 0.88 (0.78 - 0.99) |

### Supplemental Table 5

| Month | Age group | Excess Deaths (95%) CI | | | Ratio of Observed/Expected (95% CI) | | |
|---|---|---|---|---|---|---|---|
| | | All Causes | Medical | External | All Causes | Medical | External |
| 202003 | 5-9 years | -48 (-70 - -25) | -15 (-29 - -1) | -33 (-50 - -15) | 0.78 (0.71 - 0.87) | 0.81 (0.68 - 0.99) | 0.77 (0.69 - 0.88) |
| 202004 | 5-9 years | -16 (-38 - 6) | 2 (-12 - 16) | -18 (-35 - 0) | 0.91 (0.81 - 1.04) | 1.03 (0.85 - 1.31) | 0.84 (0.73 - 1.00) |
| 202005 | 5-9 years | -3 (-26 - 19) | 1 (-14 - 15) | -4 (-22 - 14) | 0.98 (0.88 - 1.11) | 1.01 (0.86 - 1.23) | 0.96 (0.84 - 1.14) |
| 202006 | 5-9 years | 23 (1 - 46) | 10 (-5 - 24) | 14 (-4 - 32) | 1.12 (1.00 - 1.26) | 1.09 (0.96 - 1.26) | 1.14 (0.96 - 1.41) |
| 202007 | 5-9 years | -1 (-23 - 21) | 1 (-13 - 15) | -2 (-20 - 15) | 1.00 (0.91 - 1.11) | 1.01 (0.90 - 1.15) | 0.98 (0.84 - 1.17) |
| 202008 | 5-9 years | 3 (-20 - 25) | 2 (-12 - 16) | 1 (-17 - 19) | 1.01 (0.91 - 1.15) | 1.02 (0.87 - 1.24) | 1.01 (0.86 - 1.21) |
| 202009 | 5-9 years | -6 (-29 - 16) | 10 (-5 - 24) | -16 (-34 - 2) | 0.96 (0.85 - 1.11) | 1.14 (0.94 - 1.43) | 0.85 (0.72 - 1.02) |
| 202010 | 5-9 years | 4 (-19 - 27) | 24 (9 - 38) | -20 (-38 - -2) | 1.02 (0.91 - 1.17) | 1.40 (1.13 - 1.86) | 0.84 (0.73 - 0.98) |
| 202011 | 5-9 years | -4 (-27 - 19) | -5 (-19 - 9) | 1 (-17 - 19) | 0.98 (0.87 - 1.12) | 0.92 (0.75 - 1.19) | 1.01 (0.87 - 1.20) |
| 202012 | 5-9 years | -55 (-78 - -32) | -5 (-20 - 9) | -50 (-68 - -32) | 0.73 (0.66 - 0.82) | 0.92 (0.76 - 1.17) | 0.64 (0.57 - 0.74) |
| 202101 | 5-9 years | -24 (-46 - -2) | 12 (-2 - 26) | -36 (-54 - -19) | 0.88 (0.79 - 0.99) | 1.19 (0.97 - 1.54) | 0.74 (0.66 - 0.85) |
| 202102 | 5-9 years | -6 (-29 - 16) | 17 (3 - 32) | -24 (-42 - -6) | 0.96 (0.85 - 1.11) | 1.32 (1.04 - 1.78) | 0.80 (0.69 - 0.94) |
| 202103 | 5-9 years | -29 (-51 - -6) | 13 (-1 - 27) | -41 (-59 - -24) | 0.87 (0.79 - 0.97) | 1.17 (0.98 - 1.44) | 0.71 (0.63 - 0.81) |
| 202104 | 5-9 years | 24 (2 - 47) | 17 (3 - 32) | 7 (-11 - 24) | 1.14 (1.01 - 1.31) | 1.26 (1.04 - 1.59) | 1.06 (0.91 - 1.28) |
| 202105 | 5-9 years | 14 (-9 - 37) | 18 (3 - 32) | -4 (-22 - 14) | 1.07 (0.96 - 1.21) | 1.22 (1.03 - 1.48) | 0.97 (0.84 - 1.14) |
| 202106 | 5-9 years | 1 (-21 - 24) | 2 (-12 - 16) | -1 (-19 - 17) | 1.01 (0.91 - 1.13) | 1.02 (0.90 - 1.17) | 0.99 (0.84 - 1.20) |
| 202107 | 5-9 years | 0 (-23 - 22) | 9 (-5 - 23) | -9 (-27 - 8) | 1.00 (0.91 - 1.11) | 1.08 (0.96 - 1.23) | 0.92 (0.79 - 1.09) |
| 202108 | 5-9 years | 47 (25 - 70) | 18 (4 - 32) | 29 (12 - 47) | 1.26 (1.12 - 1.44) | 1.22 (1.04 - 1.49) | 1.30 (1.10 - 1.58) |
| 202109 | 5-9 years | 13 (-9 - 36) | 2 (-12 - 16) | 11 (-6 - 29) | 1.08 (0.95 - 1.24) | 1.03 (0.85 - 1.28) | 1.11 (0.95 - 1.35) |
| 202110 | 5-9 years | -2 (-25 - 21) | 15 (1 - 30) | -18 (-35 - 0) | 0.99 (0.88 - 1.13) | 1.25 (1.01 - 1.63) | 0.85 (0.74 - 1.00) |
| 202111 | 5-9 years | 30 (7 - 53) | 17 (2 - 31) | 13 (-5 - 31) | 1.17 (1.03 - 1.33) | 1.26 (1.03 - 1.61) | 1.11 (0.97 - 1.31) |
| 202112 | 5-9 years | 15 (-7 - 37) | 21 (7 - 35) | -6 (-23 - 11) | 1.07 (0.97 - 1.19) | 1.30 (1.08 - 1.64) | 0.96 (0.86 - 1.09) |
| 202201 | 5-9 years | -9 (-31 - 14) | 5 (-9 - 19) | -14 (-31 - 4) | 0.96 (0.86 - 1.08) | 1.08 (0.87 - 1.43) | 0.90 (0.80 - 1.04) |
| 202202 | 5-9 years | 19 (-4 - 41) | 16 (1 - 30) | 3 (-15 - 21) | 1.11 (0.98 - 1.28) | 1.29 (1.02 - 1.76) | 1.03 (0.89 - 1.21) |

#### Supplemental Figure 4

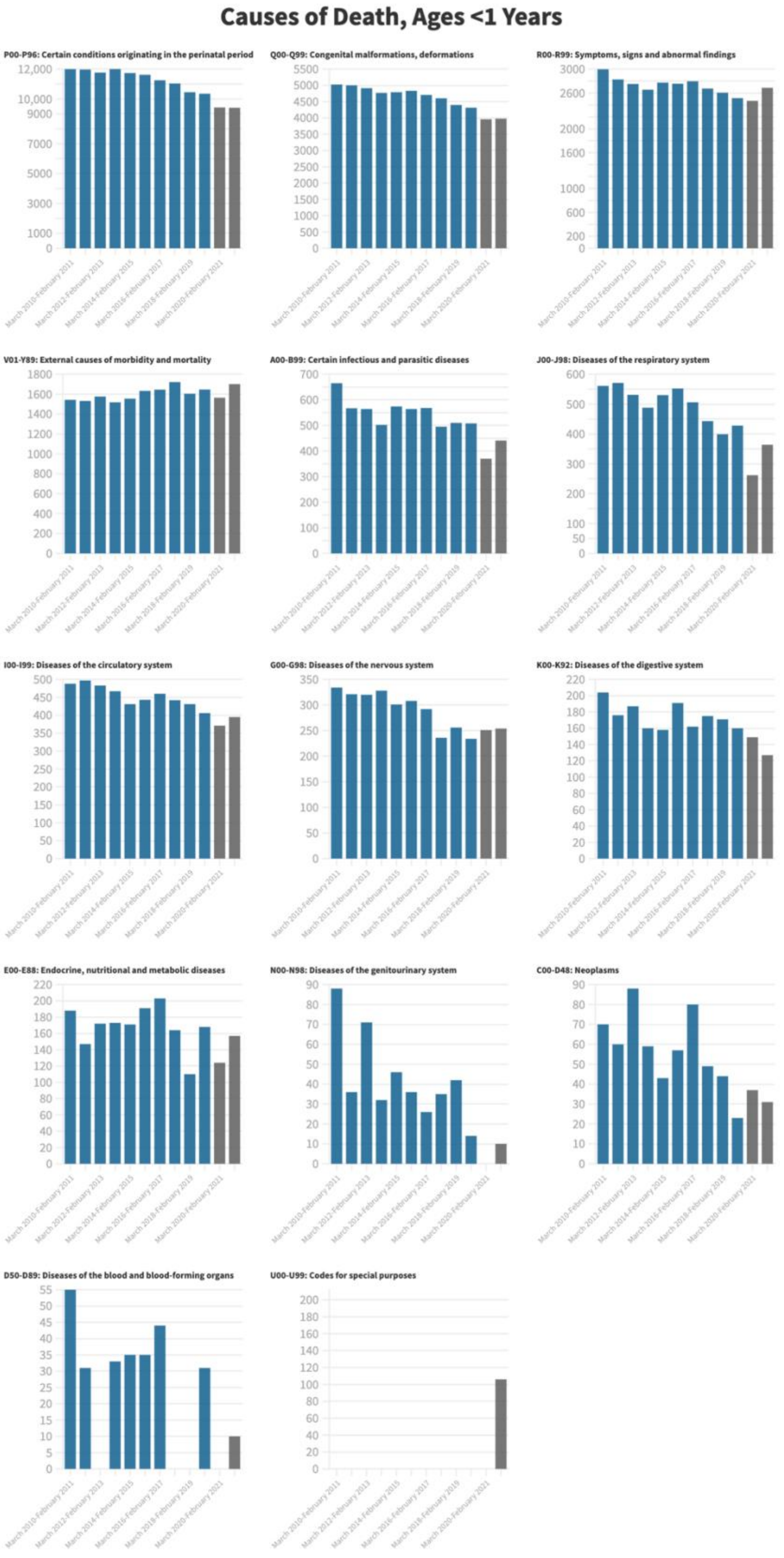


Suppression may occur for chapters with low death counts.
Note that U00-U99 includes deaths from COVID-19.

### Supplemental Figure 5

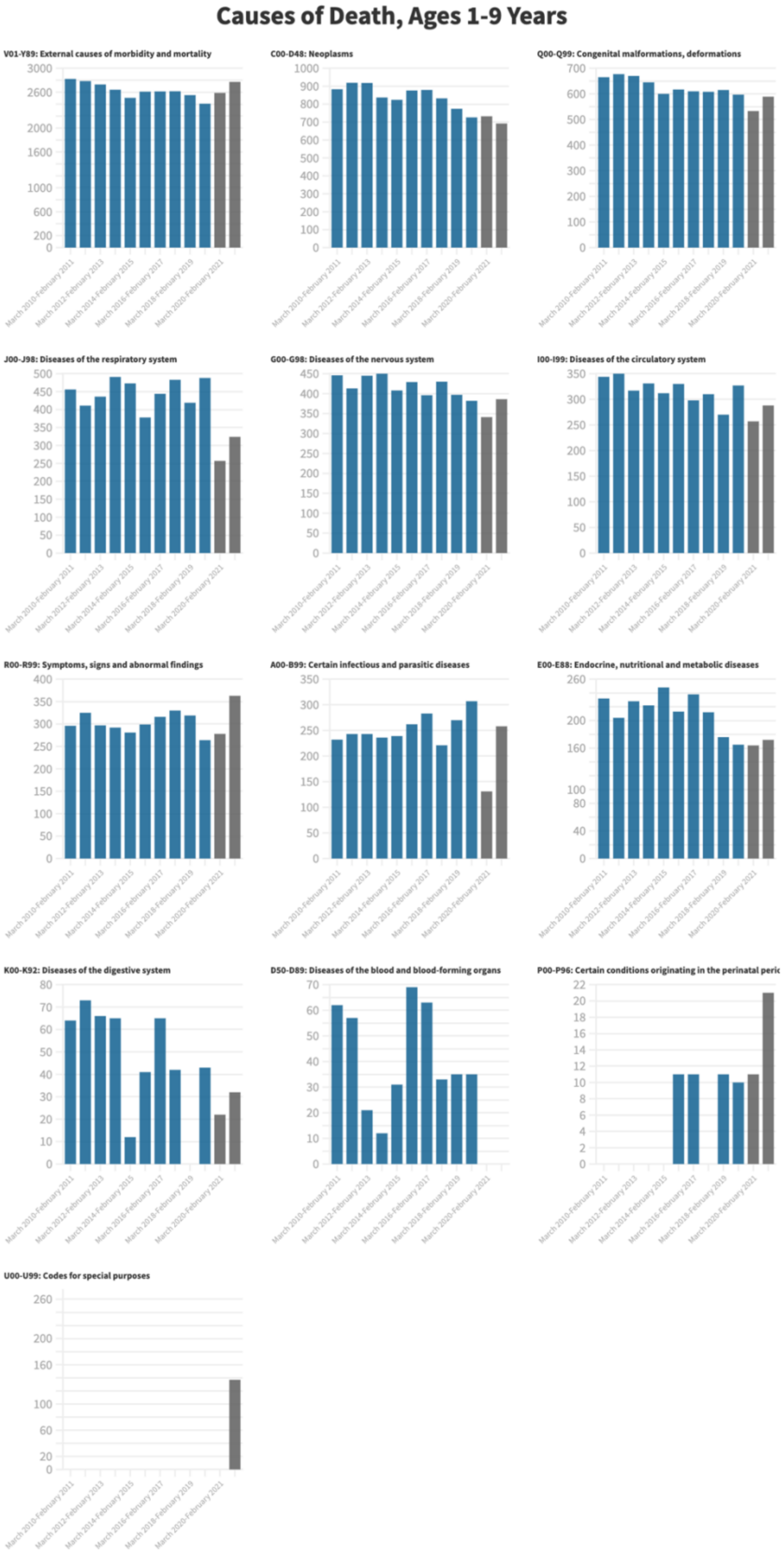


Suppression may occur for chapters with low death counts.
Note that U00-U99 includes deaths from COVID-19.

**Supplemental Figure 6**

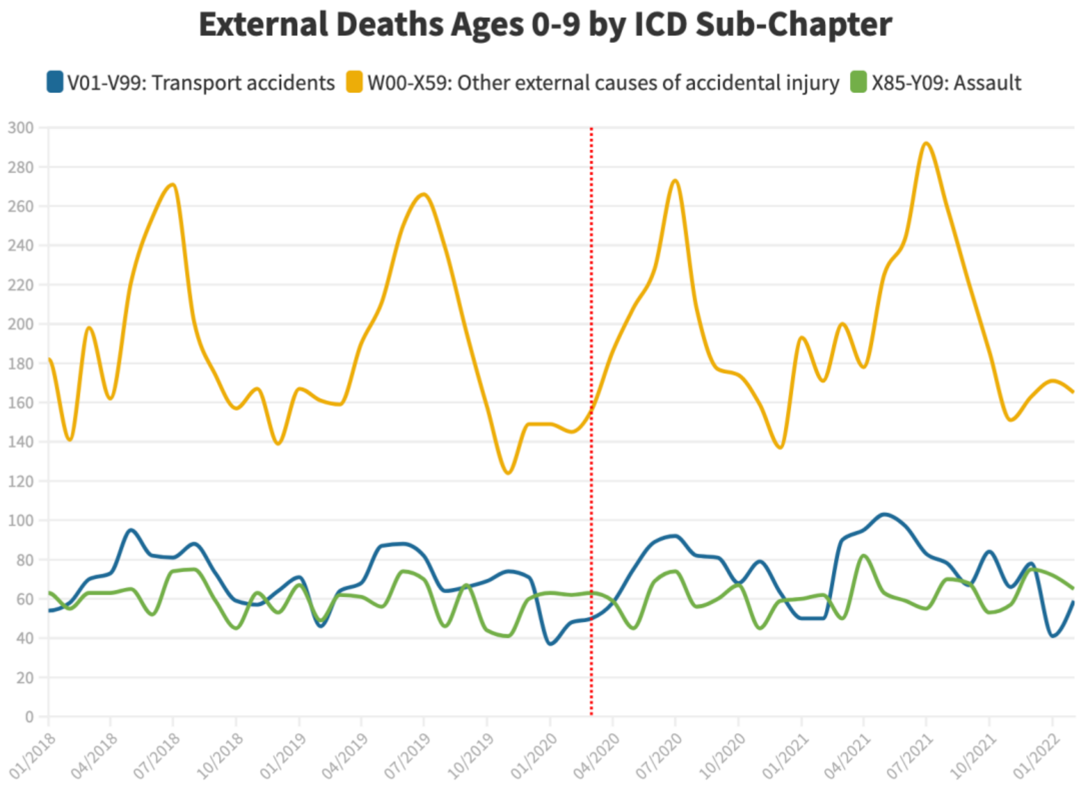
